# Prevalence and incidence of schizophrenia: Temporal and regional trends in Germany

**DOI:** 10.1101/2025.04.05.25325295

**Authors:** Oliver Riedel, Christian J. Bachmann, Robert A. Bittner, Michael Dörks, Bianca Kollhorst, Mishal Qubad, Oliver Scholle

## Abstract

**Background:** Schizophrenia ranks among the top ten causes of disability worldwide. The provision of healthcare services requires estimates on the epidemiology of schizophrenia, but recent data for Germany are lacking.

**Methods:** Based on a large German health claims database (GePaRD), we identified persons aged 0–64 years with treated schizophrenia as those having an inpatient/outpatient ICD-10 diagnosis (F20) and a prescription for a schizophrenia-recommended antipsychotic in the same calendar year. For each year from 2012 to 2021, we calculated the standardized incidence proportion (SIP) and the prevalence of schizophrenia. Analyses were stratified by sex, age, and population density in the region of residence.

**Results:** The SIP of treated schizophrenia remained stable from 2012 to 2017 (46.0– 46.5/100,000) and subsequently declined to 41.3/100,000 in 2021, with higher SIP in men (45.3/100,000) than in women (37.1/100,000). In 2021, the SIP was comparable in urban, rural, and sparsely populated rural districts (36.3–38.5/100,000) and higher in large urban cities (48.3/100,000). SIP estimates among children and adolescents (aged 0–17 years) varied between 3.5/100,000 and 4.1/100,000 over the study period. The standardized prevalence of schizophrenia declined from 366.1/100,000 in 2012 to 334.0/100,000 in 2021.

**Conclusion:** Similar to other Western countries, there has been a decline in the incidence and prevalence of schizophrenia in Germany over the last few years. The higher incidence in males and those living in large urban areas highlights the health care needs of these populations.

## INTRODUCTION

Schizophrenia is a severe mental disorder within the spectrum of psychotic disorders, characterized by positive symptoms such as hallucinations, delusions, and disorganized thinking, negative symptoms including flattened or inadequate affect and alogia as well as pervasive cognitive dysfunction. Patients are most often diagnosed in their twenties [1]. In rarer cases, the disorder can manifest during childhood, i.e. before the age of 13, or adolescence before the age of 18, commonly referred to as childhood-onset schizophrenia (COS) and early-onset schizophrenia (EOS) respectively [2]. However, it is difficult to determine the exact onset of the disorder due to the often unspecific prodromal phase, resulting in a considerable duration of untreated psychosis (DUP), during which patients are manifestly ill without being adequately diagnosed and treated with antipsychotics [3].

Systematic reviews have estimated a lifetime prevalence of schizophrenia between 4.8 and 7.2 per 1,000 inhabitants [4, 5]. The Global Burden of Disease study reported an estimated prevalence of schizophrenia of 287/100,000 and an incidence of 16/100,000 in 2019, with mildly decreasing values of around 1 to 3 percent for both measures since 1990 and a stable male-female-ration of 1.1 [6]. Findings also suggest that both prevalence and incidence of schizophrenia are associated with urbanicity as well as sociodemographic indices, with higher frequencies in populations of higher urbanicity and low socioeconomic status [7, 8].

Although the frequency of schizophrenia is low compared with other mental disorders, it nonetheless has a substantial impact on public health; it also creates a substantial socioeconomic burden for patients as well as their families and caregivers. The former is reflected in substantially reduced life expectancy with years of potential life lost (YPLL) globally estimated at 15.2 years [9]. Moreover, globally age-standardized disability-adjusted life years (DALYs) have been estimated at 184 per 100,000. For Europe, the direct annual costs of illness have been estimated at 29 billion euros [10, 11]. Apart from the individual cases of illness, a precise assessment of the frequency of the disorder is therefore of great importance from a socioeconomic perspective in order to estimate the need for treatment and optimize the provision of healthcare services.

The aim of this study was to assess and characterize the incidence and prevalence of schizophrenia in Germany in children, adolescents, and adults across temporal and regional dimensions in a real-world setting and to describe those by urbanicity, socioeconomic status and sex.

## METHODS

We conducted a population-based cross-sectional study, analyzing routinely collected German healthcare data.

### Data source

We used the German Pharmacoepidemiological Research Database (GePaRD), which is based on claims data from four statutory health insurance providers in Germany and currently includes information on approximately 25 million persons who have been insured with one of the participating providers since 2004 or later [12]. Per data year, GePaRD covers approximately 20% of the general population and all geographical regions of Germany are represented. Demographic data include sex, age, and region of residence on the district level. Prescription data in GePaRD include all reimbursed drugs prescribed by general practitioners or specialists in the outpatient setting. Prescriptions are coded according to the German modification of the WHO Anatomical Therapeutic Chemical (ATC) classification system (version from April 2023 for this study). In addition, GePaRD contains information on outpatient (i.e., from general practitioners and specialists) and inpatient services and diagnoses. Diagnoses are coded according to the German modification of the International Classification of Diseases and Related Health Problems, 10th Revision (ICD-10-GM).

### Study design

To be eligible for the year-wise study populations from 2012 to 2021, each person had to have (a) valid information on sex and an age between 0 and 64 years, (b) German residency, and (c) continuous insurance coverage in the respective year (gaps of up to 30 days were allowed). Persons who died or were born in the respective year were included but only required to be continuously insured from January 1 or until December 31, respectively. For estimating the incidence, eligible persons additionally had to have both continuous insurance for the two years prior to the respective year and no history of treated schizophrenia (definition see below) during these two years.

### Identification of persons with schizophrenia

To enhance the validity of identifying individuals with schizophrenia, we used a more stringent approach than relying solely on coded diagnoses, by focusing on persons with treated schizophrenia. A person was considered as treated for schizophrenia in a given year, if they had an in- or outpatient diagnosis of schizophrenia (ICD-10-GM code F20) in conjunction with at least one outpatient prescription of an antipsychotic considered potentially relevant for the treatment of schizophrenia (see **Suppl. Table S1**).

In addition to this main definition, we conducted two sensitivity analyses deviating from the primary approach by the following aspects. The first analysis considered only inpatient diagnoses of schizophrenia. In the second analysis, we included a broader range of diagnostic codes (both inpatient and outpatient) comprising schizophrenia spectrum disorders (ICD-10-GM codes F20, F21, F22, F23, F25, F28, and F29).

### Regional characteristics

Via the person’s region of residence, we linked district-level characteristics (total number of districts: 401) including urbanicity and socioeconomic deprivation. Urbanicity was defined by the type of district according to settlement structure (as of 2017), categorized into four classes ranging from “large urban city” to “sparsely populated rural district”. Socioeconomic deprivation was measured using the German Index of Socioeconomic Deprivation (GISD; as of 2018). This index, developed by the Robert Koch Institute, quantifies regional socioeconomic deprivation in Germany by integrating indicators of education, employment, and income (13).

### Data analysis

Separately for each year, we calculated the incidence proportion of schizophrenia, defined as the number of persons with newly identified schizophrenia who had not been identified as having schizophrenia in the two years before, per population (expressed as “per 100,000 persons”). We also calculated the prevalence of schizophrenia per population following the same methodology but omitting the two-year pre-observation period requirement.

Calculations were made overall and stratified by sex, age groups, and regional characteristics (only for the incidence). Persons younger than 18 years were considered for estimating EOS (13-17 years) and COS (younger than 13 years), respectively. Prevalence and incidence estimates were calculated along with 95% confidence intervals (CIs) and directly standardized according to age and sex with reference to the German population as of December 31, 2021. All statistical analyses were conducted using SAS version 9.4 (SAS Institute, Cary, NC, USA).

## RESULTS

### Incidence overall and by sex and age

The number of eligible persons for estimating the incidence ranged between 9,589,084 (2012) and 12,450,531 (2021) (**Suppl. Table S2**). In 2021, the standardized incidence proportion was 41.3/100,000 (95% CI: 40.1–42.4) overall and 45.3/100,000 (95% CI: 43.6–47.0) in males and 37.1/100,000 (95% CI: 35.6–38.6) in females.

The standardized incidence in 2021 by sex and age is shown in **Figure 1**. In males, incidence proportions were highest in the age groups between 20 and 25 years (76.2–80.2/100,000); in females, the incidence proportions past the age of 18 years were similar across the age groups. When considering only inpatient diagnoses of schizophrenia, lower incidence proportions— e.g., 14.4/100,000 (95% CI: 13.7–15.0) overall in 2021—but similar age patterns were observed (see **Suppl. Figure S1**).

**Figure 1:**
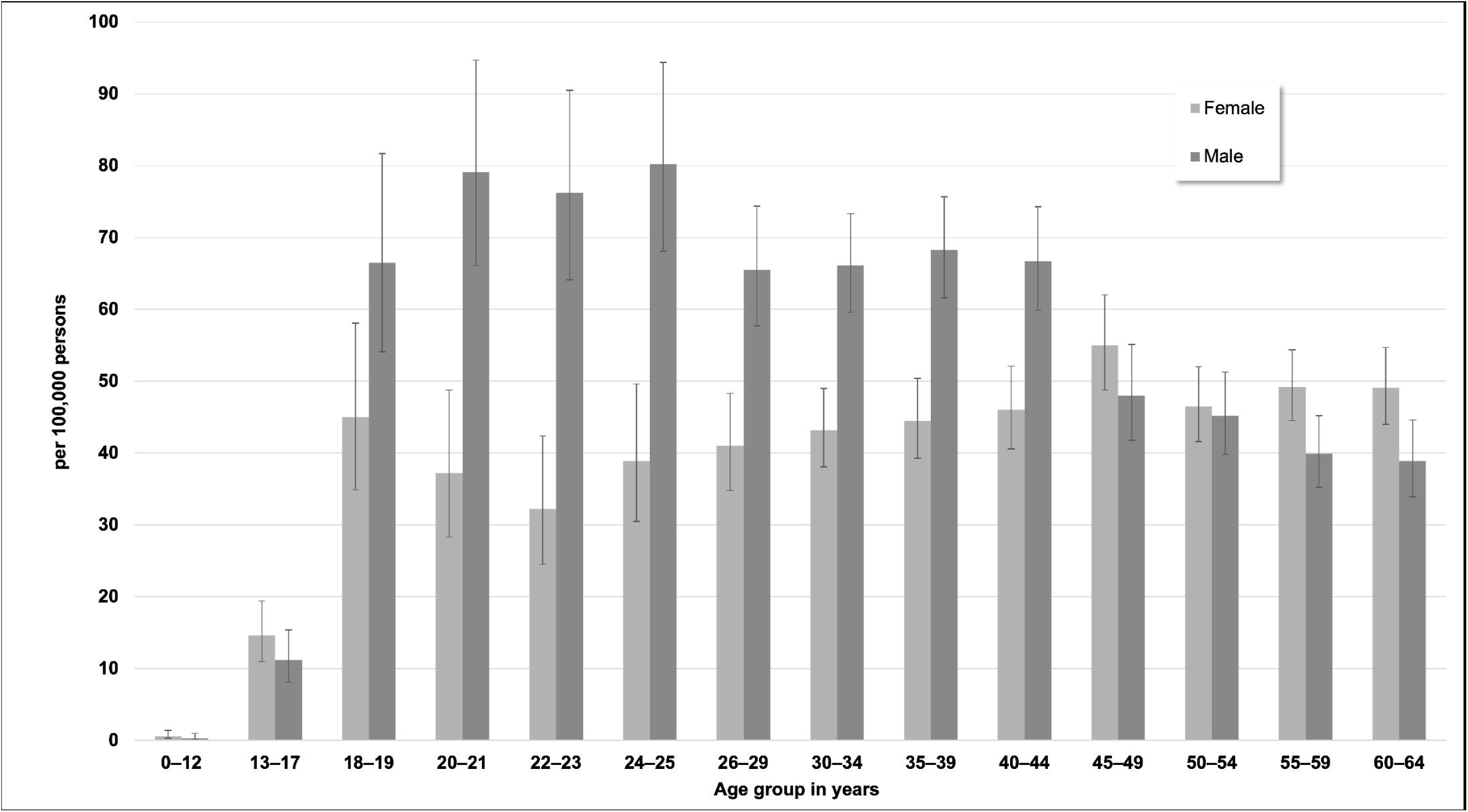
Standardized incidence proportions (with 95% CIs) of schizophrenia by sex and age in 2021

The development of the incidence of schizophrenia during the study period is shown in **Figure 2**. Overall, the incidence remained stable from 2012 (46.0/100,000, 95% CI: 44.6–47.4) to 2017 (46.5/100,000, 95% CI: 45.2–47.7), followed by a subsequent decline to 41.3/100,000 (95% CI: 40.1–42.4) in 2021. Age- and sex-wise comparisons of the incidences between 2012 and 2021 indicated that the decline over the years was driven by a decline in the age groups 26–34 years among males and 30–64 years among females (see **Suppl. Figure S2**).

**Figure 2:**
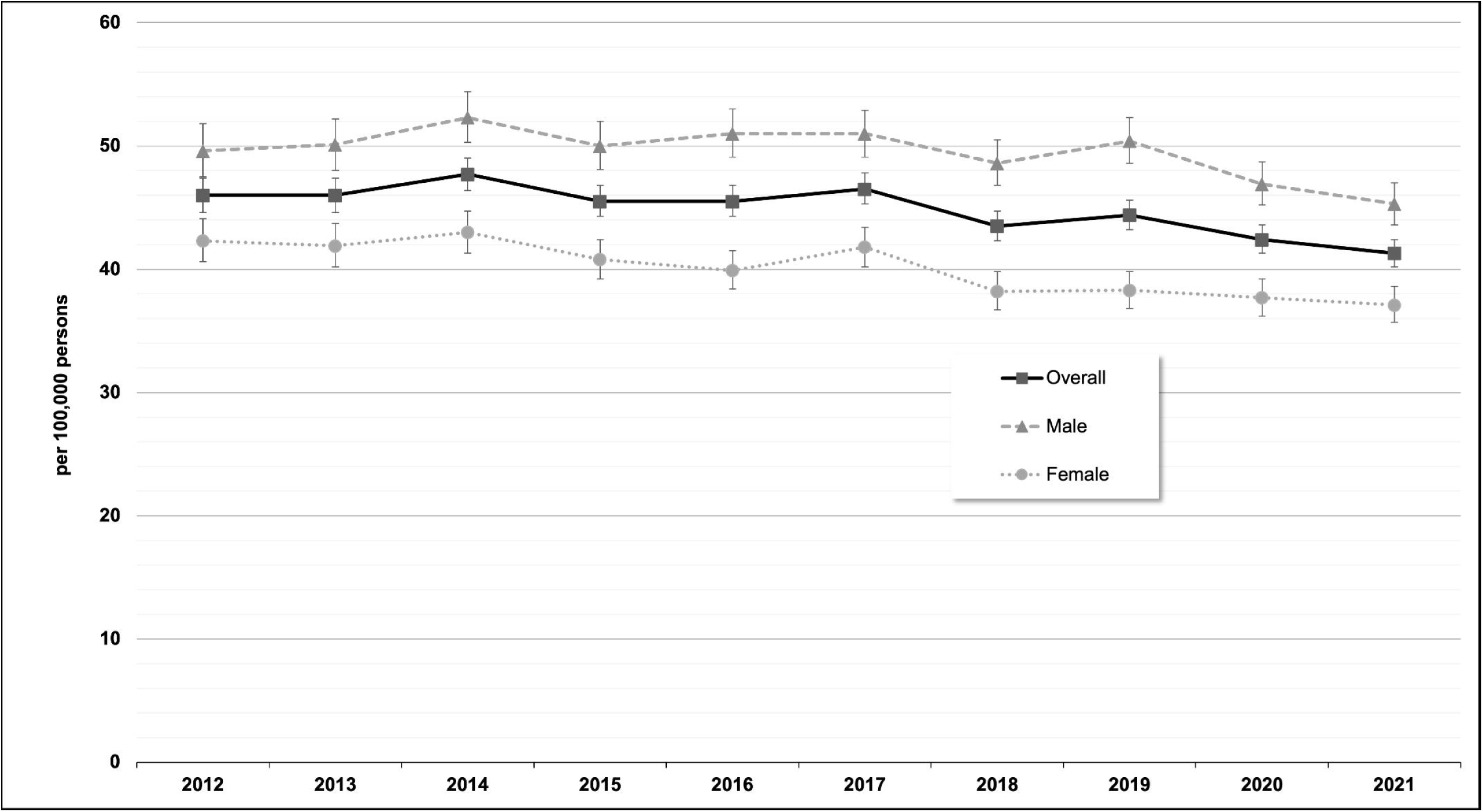
Standardized incidence proportions (with 95% CIs) of schizophrenia by sex and calendar year

The first sensitivity analysis, which considered only inpatient schizophrenia diagnoses, showed a similar development over time, albeit at a lower level (as mentioned above in 2021: 14.4/100,000, 95% CI: 13.7–15.0; see **Suppl. Figure S3**). The second sensitivity analysis using a broader range of schizophrenia spectrum disorders also showed a similar development over time but with higher incidence proportions (2021: 64.4/100,000, 95% CI: 63.0–65.8; see Suppl. Figure S3).

Regarding EOS, the overall incidence proportions varied between 12.0/100,000 (95% CI: 9.7–14.9) in 2014 and 14.6/100,000 (95% CI: 11.9–17.8) in 2019, with similar estimates for males and females and no clear temporal trend (see **Figure 3**). For COS, no estimates are shown due to the low number of affected children (between 2 and 9 children per data year).

**Figure 3:**
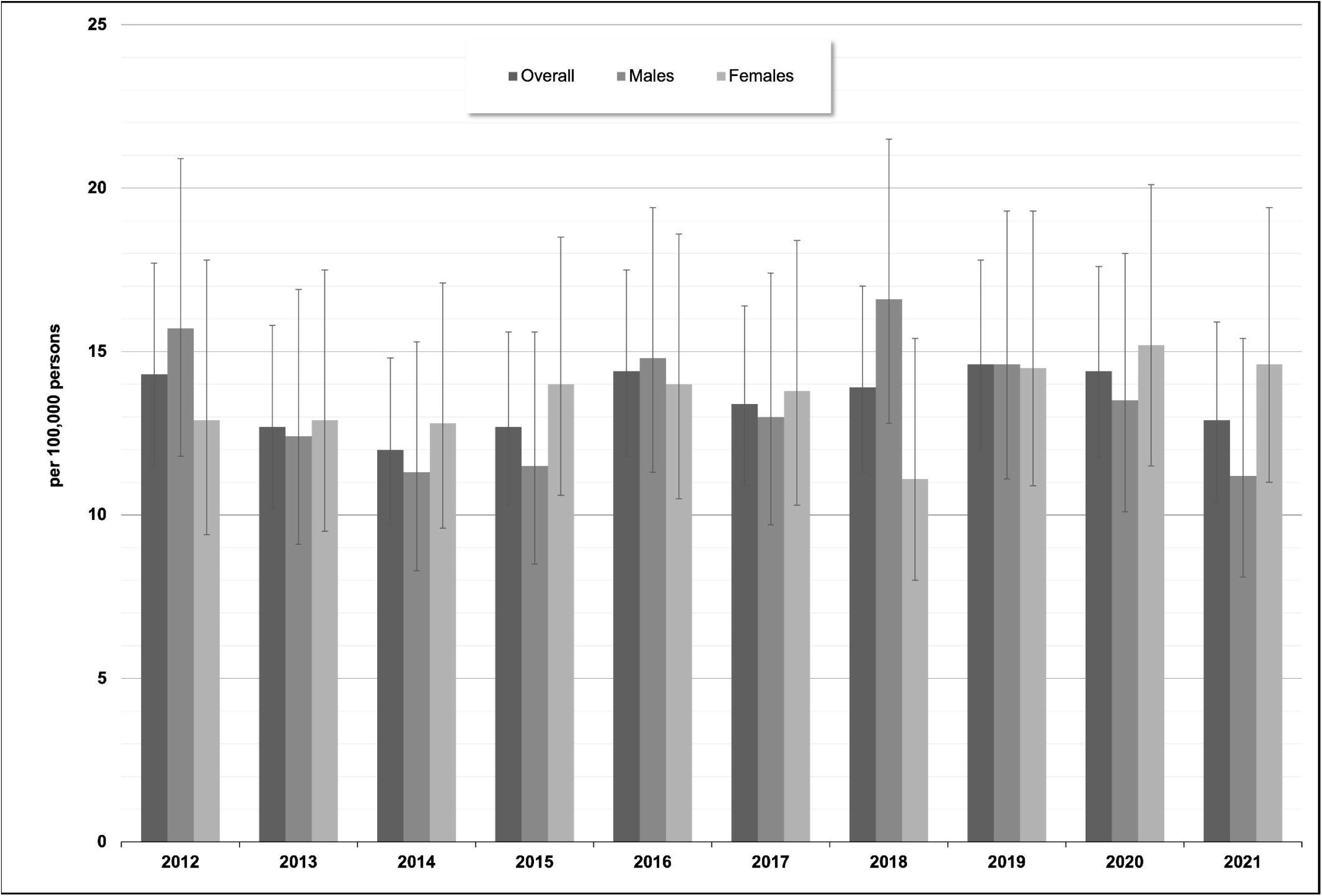
Standardized incidence proportions (with 95% CIs) of early-onset schizophrenia (age group 13–17 years) by sex and calendar year

### Incidence by regional characteristics

Analyses stratified by urbanicity revealed a higher standardized incidence proportion of schizophrenia in large urban cities (48.3/100,000; 95% CI: 46.2–50.4) compared to urban districts (38.5/100,000; 95% CI: 36.7–40.3), rural districts (37.9/100,000; 95% CI: 35.1–41.0), and sparsely populated rural districts (36.3/100,000; 95% CI: 33.3–39.6) in 2021 (see **Figure 4**). These differences in schizophrenia incidence were more pronounced in 2012 compared to 2021, primarily due to a 17% decrease in the incidence proportion in large urban cities over this period (from 58.5 to 48.3/100,000), along with unchanged or less pronounced decreases in the other classes of urbanicity.

**Figure 4:**
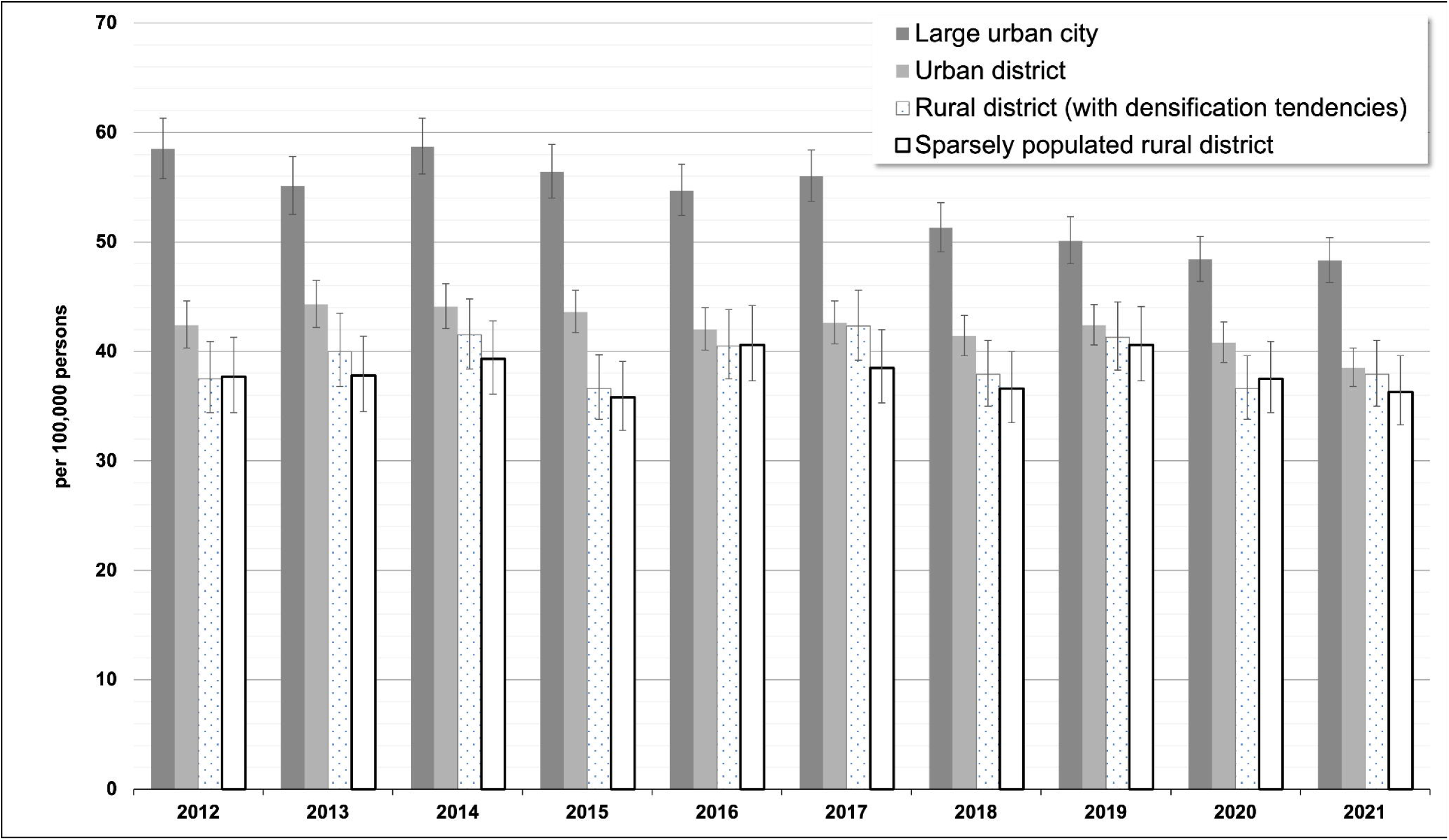
Standardized incidence proportions (with 95% CIs) of schizophrenia by urbanicity and calendar year

The standardized incidence proportion of schizophrenia also differed across regional socioeconomic strata. In 2021, the 20% most deprived districts had a higher incidence of schizophrenia (48.6/100,000) compared to less deprived areas (60% moderately deprived districts: 40.8/100,000; 20% least deprived districts: 39.5/100,000; see **Suppl. Figure S4**). The observed difference in 2021 developed over the course of the study period. Until the end of 2015, differences were minimal. From 2019 to 2021, however, schizophrenia incidence increased in 20% of the districts, namely those with the highest level of socioeconomic deprivation, while a decrease was seen in the two classes encompassing less deprived districts.

### Prevalence

The number of eligible persons for estimating the prevalence ranged between 11,653,946 (2012) and 13,499,924 (2021). The overall prevalence of schizophrenia declined by 9% from 366.1/100,000 (95% CI: 362.6–369.6) in 2012 to 334.0/100,000 (95% CI: 331.0–337.1) in 2021, with higher prevalences in males than in females in each year (see **Figure 5**). From 2012 to 2021, the relative decrease in the prevalence of schizophrenia was about twofold higher in women (−13%) compared to men (−5%).

**Figure 5:**
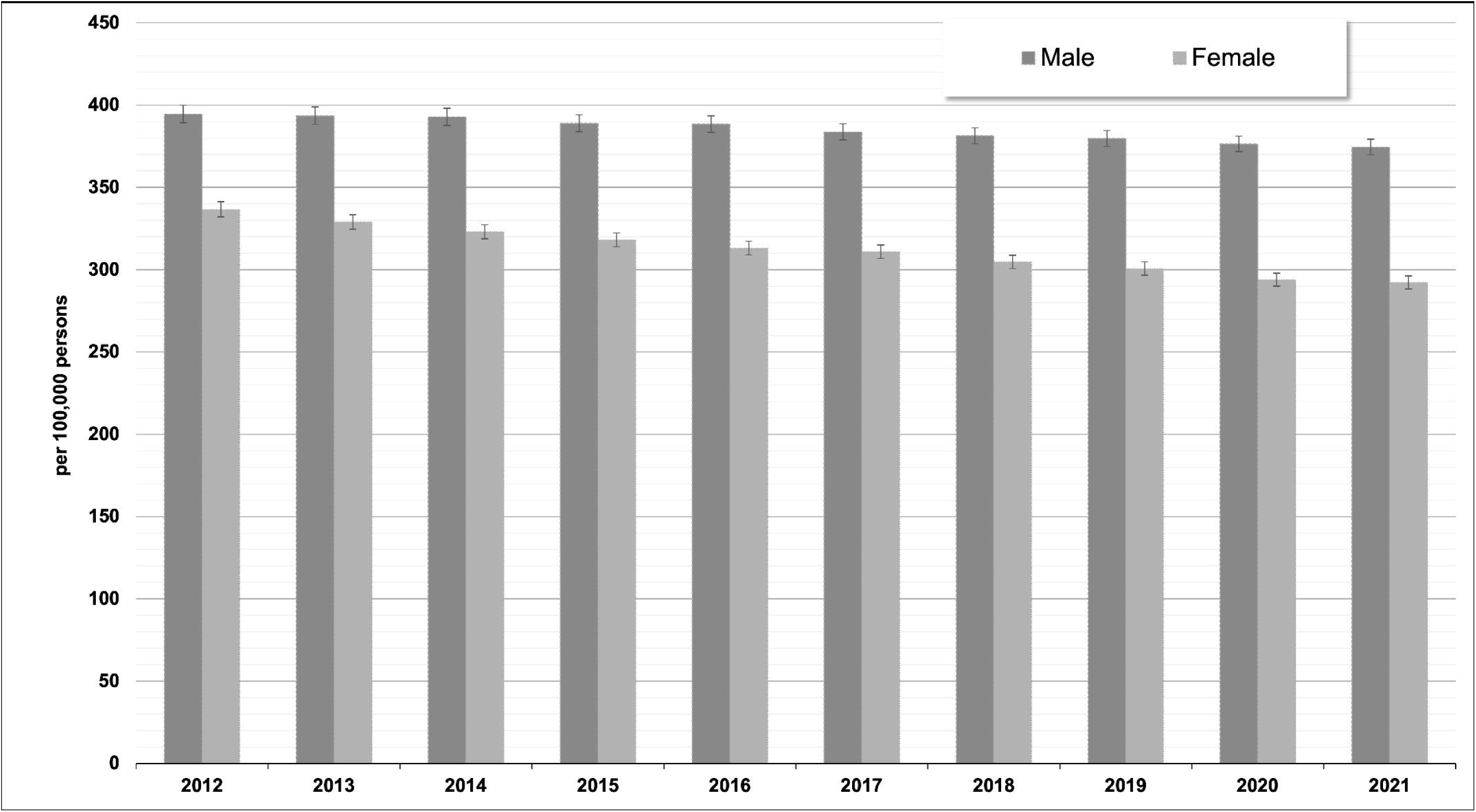
Standardized prevalence (with 95% CIs) of schizophrenia by sex and calendar year

## DISCUSSION

We investigated the incidence and prevalence of schizophrenia from 2012 to 2021 in Germany and assessed differences by age, sex, and urban density of the region based upon health claims data. In 2021, we found an overall age- and sex-standardized incidence of schizophrenia of 41.3/100,000 when considering both outpatient and inpatient diagnoses. When including inpatient diagnoses only, the incidence decreased to 14.4/100,000.

The incidences of both definitions are consistent with epidemiological estimates that have been previously published. Based on systematic reviews, McGrath et al. [4] reported a median incidence of 15.2/100,000, with 80% of estimates ranging between 7.7 and 43/100,000. Solmi et al. [6] estimated an incidence for schizophrenia of 16.9/100,000 in 1990 and 16.3/100,000 in 2019, constituting a relative decline of 3.3% during that span. Of note, a decline was also seen in our data from 2012 to 2021, though with higher relative declines of 10% for the main definition and 22% for the inpatient definition.

Regarding the age- and sex-specific patterns of the incidences, we found highest incidence rates in men between the age of 20 and 25 years, whereas in women incidence rates were lower and largely comparable across age groups over the age of 18 years. These sex-specific differences in the evolution of incidence rates across age groups are well known. For example, our data resemble findings of a Finnish cohort study, which also reported a decline of incidences in men past the end of their twenties while in women the incidences are largely comparable across these age groups [14]. These findings dovetail with a recently published meta-analysis of 192 studies on the age of onset of mental disorders, which estimated a median age of onset of 25 years with an interquartile range of 21 to 35 years for schizophrenia [1].

The large disparities between the incidences according to our main definition and the definition restricted to inpatient diagnoses might be surprising, assuming that most patients with incident schizophrenia initially require hospitalization. One potential explanation relates to the structure of German health care, which, among others, offers treatment conducted in specialized psychiatric facilities associated with hospitals, yet classified as outpatient facilities in the claims data. Nevertheless, we also assume that the disparities between the estimates can be attributed, at least in part, to the inclusion of less severe cases of schizophrenia in the main analysis due to the consideration of outpatient diagnoses. However, the incidence estimates from both definitions remain within the range of epidemiological estimates (as described above), and our objective was to estimate the frequency of treated schizophrenia in a real-world setting.

Among children and adolescents, the incidence of COS/EOS was between 3.3 and 4.1/100,000 in our study. This figure was lower than a previously reported incidence of 9/100,000 as estimated in a French cross-sectional assessment [15]. Their findings, however, have to be treated with caution due to methodological limitations regarding the low participation quota of patients and study centers. Regarding the overall incidence of EOS (i.e. among those aged 13–17 years), our findings are largely comparable to those presented by Okkels et al. [16]. Based on the Danish Psychiatric Central Research Register (DPCRR), which includes data from all admissions to psychiatric hospitals in Denmark since 1969) the authors estimated an incidence rate of 16/100,000 among those aged 12–18 years for the period of 1994 to 2010. However, there is still a considerable variance with regard to the estimated incidence of schizophrenia in younger age groups. Driver et al. [2] found an incidence of COS of 40/100,000 based on an NIMH study and considering patients below the age of 13 years. Recently published findings from a Finnish register study even suggest an incidence of COS/EOS up to 70/100,000 in females and 20/100,000 in males below the age of 18 years [17]. Regarding these heterogeneous results, our own findings are difficult to interpret. It must also be borne in mind that, particularly in younger age groups, there is often a considerable symptomatic overlap with other mental disorders (e.g., autism spectrum disorders, developmental delay), which makes an exact diagnosis even more difficult [18–20]. In addition to that, Vernal et al. [21] reported based on a register study that outpatient diagnoses of EOS can be of lower validity, which might also have contributed to our results. Furthermore, the DUP in patients with COS/EOS is considerably longer than in adult-onset schizophrenia (average DUP = 18.7 months in COS/EOS vs. 10.7 months in adult-onset schizophrenia [3]. This discrepancy might have contributed to an underestimation of COS/EOS cases.

We found that the incidence of schizophrenia varied by urbanicity and regional socioeconomic strata. The higher incidence of schizophrenia in large urban cities aligns with prior research, which showed that individuals in the most urbanized environments exhibited more than double the risk of schizophrenia compared to those in the most rural areas [8]. Potential explanations for this association include environmental and social stressors, such as social fragmentation, material deprivation, and increased exposure to air pollution, all of which could contribute to heightened stress [22, 23]. Another possible explanation is that stigmas associated with mental illness may discourage individuals from seeking care in rural areas, or that rural physicians may be hesitant to formally code schizophrenia due to concerns about labeling individuals in a socially restrictive environment. The observed higher incidence of schizophrenia in districts with lower socioeconomic status supports previous findings that socioeconomic disadvantage is associated with an increased risk of schizophrenia [7, 24].

We found an overall age- and sex-standardized prevalence of 366/100,000 in 2012 and 334/100,000 in 2021, respectively, both of which are in line with previous findings. An early meta-analysis based on more than 150 studies reported a slightly higher prevalence rate of 460/100,000 (4). Simeone et al. [5] found a median 12-month-prevalence of 330/100,000 (interquartile range: 260/100,000–510/100,000), based on a systematic review, while the earlier mentioned GBD study estimated a prevalence of 287/100,000 in 2019 [6]. As was the case for the incidence estimates, we also found a decline in the prevalence estimates of around 9% from 2010 to 2021, which again was higher than the decline as reported from the GDB study data (0.9%). Similarly, a recently published study on the prevalence of diagnostic codes for mental disorders reported a relative decline of 12% for schizophrenia (ICD-10 F20-F29) from 2012 to 2022 [25]. However, these findings, though also based on German health claims data, have to be interpreted with caution, since the underlying study already counted the single occurrence of a code, while the algorithm established in our analyses was stricter to increase validity.

### Strengths and limitations

The main strength of the present study is the underlying health claims data base, which comprised data from a large proportion of the German population over a comparably long period of time. Both aspects enabled us to study temporal and regional trends of schizophrenia based on a large sample. Moreover, our findings potentially contribute to research on the epidemiology of EOS, which still can be considered understudied. Our chosen case definition of schizophrenia, which was based on in- and outpatient diagnoses in conjunction with specific prescriptions can also be regarded as highly valid. In fact, the consistency of both the estimated frequency of schizophrenia and the observed differences by urbanicity and district-level socioeconomic status with findings from prior epidemiological studies suggest that GePaRD is feasible for schizophrenia research.

However, in addition to the previously mentioned limitations, further flaws should be kept in mind when interpreting our findings. For example, as our case definition was based on ICD-10 diagnoses, persons with psychotic symptoms yet below the threshold for a final diagnosis of schizophrenia were not considered in our analyses as these cases cannot be identified in health claims data. A further limitation is the lack of information regarding migration status, which has been previously identified as a risk factor [26, 27].

In conclusion, even when considering the slight declines in the frequencies of schizophrenia over the last years, our real-world data still underline the public-health burden associated with this psychiatric disorder. This is especially true for male patients and those living in large urban areas, which underscores the need for optimized care structures for these populations.

## SUPPLEMENTARY MATERIAL

For supplementary material accompanying this paper, visit cambridge.org/EPA.

## ACKNOWLEDGEMENTS

The authors would like to thank all statutory health insurance providers which provided data for this study, namely AOK Bremen/Bremerhaven, DAK-Gesundheit, Techniker Krankenkasse (TK), and hkk Krankenkasse. They would also like to thank Alina Ludewig and Fabian Gesing for the statistical programming of the data and Dr. Heike Gerds for proofreading the final manuscript (all with Leibniz Institute – BIPS).

## AUTHOR CONTRIBUTIONS

OR wrote the first draft of the manuscript, OS reviewed the manuscript and provided critique and conceptualized the study design, BK conducted all statistical analyses and reviewed the manuscript, CB, MD, RAB and BK assisted in conceptualizing the study design, helped with data interpretation, and critically reviewed the manuscript. MQ helped with data interpretation and critically reviewed the manuscript.

## ETHICS STATEMENT

In Germany, the utilization of health insurance data for scientific research is regulated by the Code of Social Law. All involved health insurance providers as well as the Federal Office for Social Security and the Senator for Health, Women and Consumer Protection in Bremen as their responsible authorities approved the use of GePaRD data for this study. Informed consent for studies based on claims data is required by law unless obtaining consent appears unacceptable and would bias results, which was the case in this study. According to the Ethics Committee of the University of Bremen studies based on GePaRD are exempt from institutional review board review.

## DATA AVAILABILITY STATEMENT

As we are not the owners of the data, we are not legally entitled to grant access to the data of the German Pharmacoepidemiological Research Database. In accordance with German data protection regulations, access to the data is granted only to employees of the Leibniz Institute for Prevention Research and Epidemiology – BIPS on the BIPS premises and in the context of approved research projects. Third parties may only access the data in cooperation with BIPS and after signing an agreement for guest researchers at BIPS.

## SUPPLEMENTARY MATERIAL (LEGENDS)

**Table S1:** Antipsychotics used in this study for the identification of persons with treated schizophrenia

**Table S2:** Standardized incidence proportion (with 95% CIs) of schizophrenia by demographic characteristics and calendar year (per 100,000 persons)

**Figure S1:**
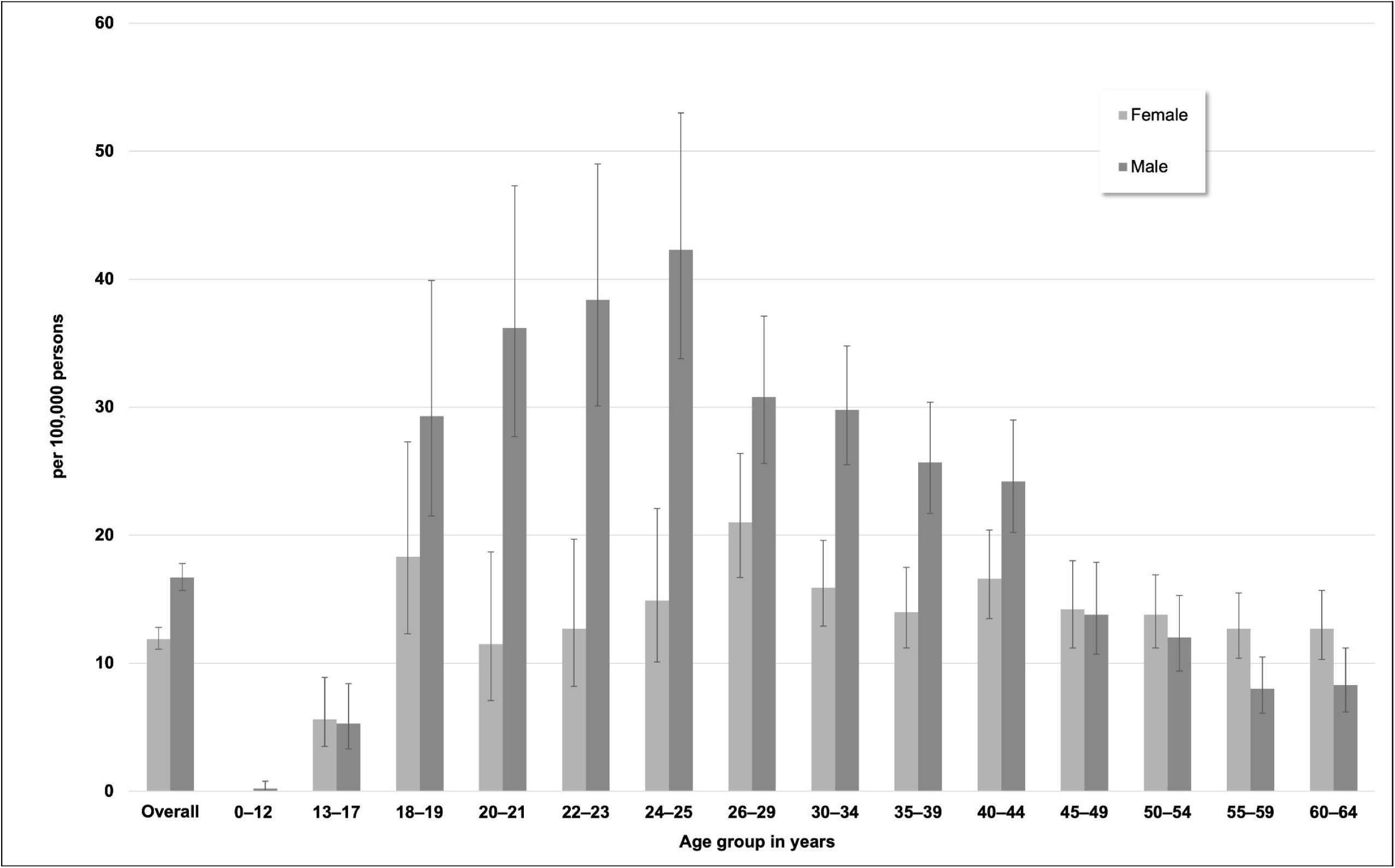
Standardized incidence proportions of schizophrenia (with 95% CIs) by sex and age in 2021 when considering only inpatient diagnoses

**Figure S2:**
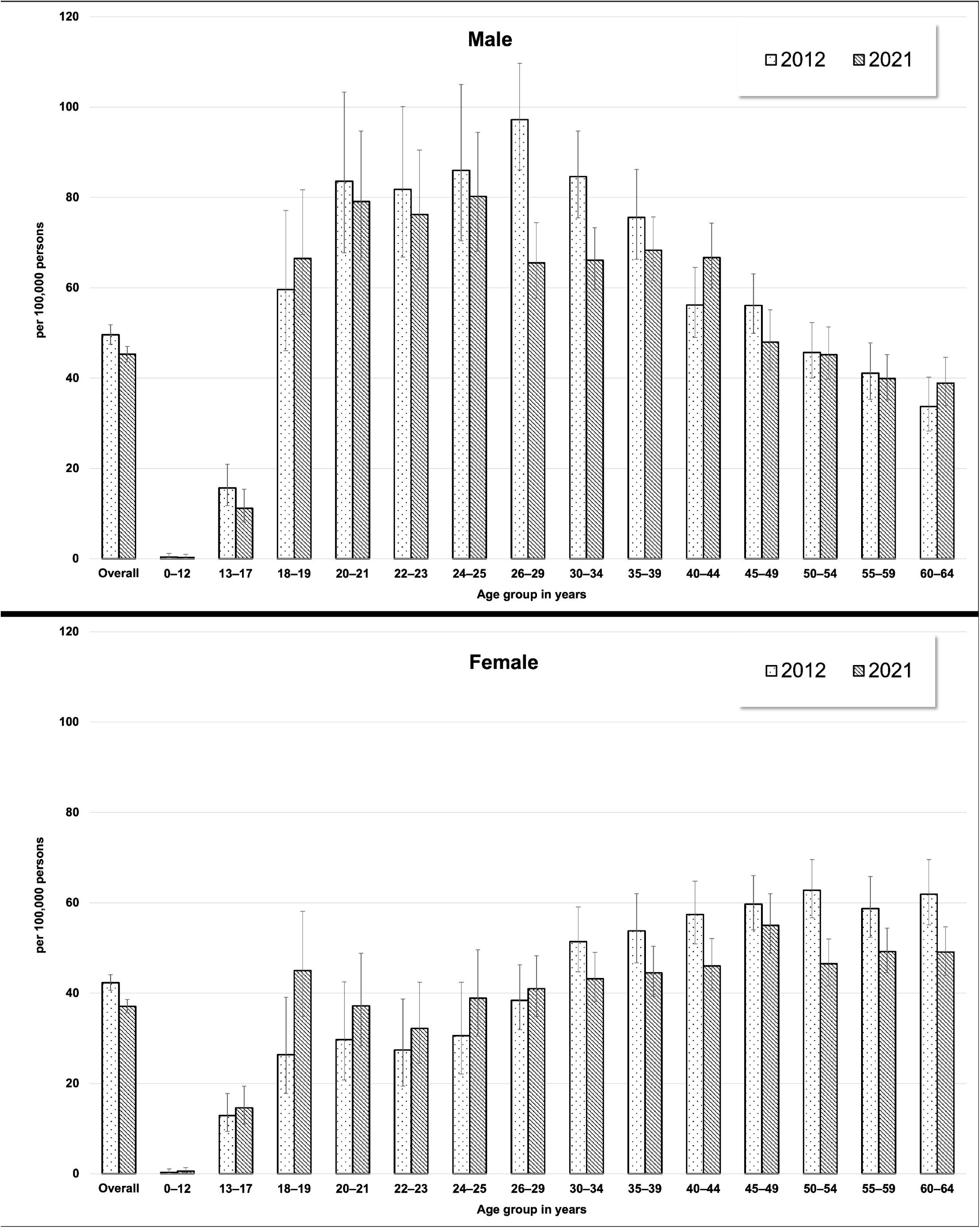
Standardized incidence proportions (with 95% CIs) of schizophrenia in 2012 and 2021 by age among males (upper figure) and females (lower figure)

**Figure S3:**
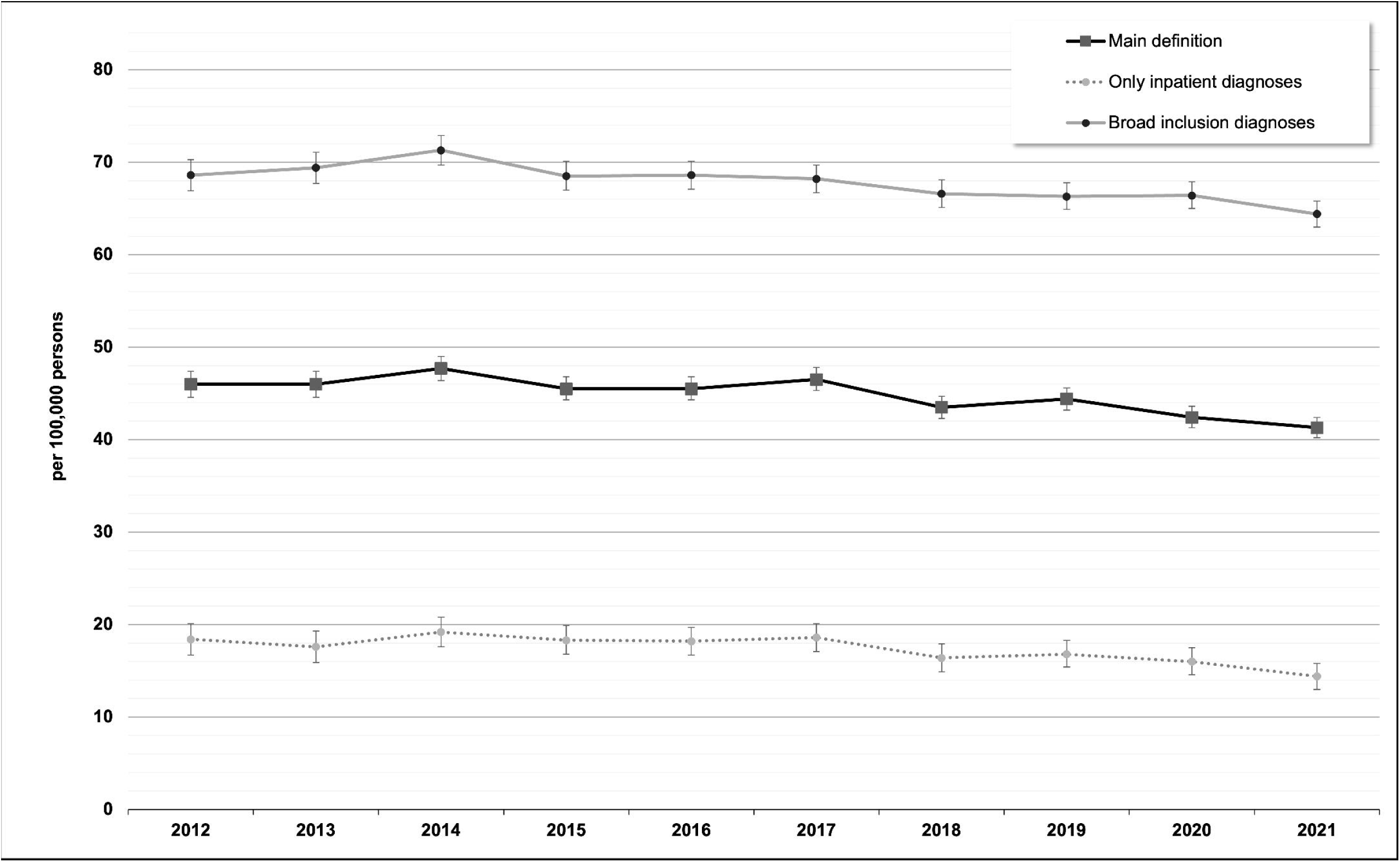
Overall standardized incidence proportion of schizophrenia (with 95% CIs) by calendar year according to (a) the main definition (antipsychotic plus outpatient or inpatient diagnosis F20), (b) when considering only inpatient diagnoses, or (c) when considering broad inclusion diagnoses (F20, F21, F22, F23, F25, F28, F29)

**Figure S4:**
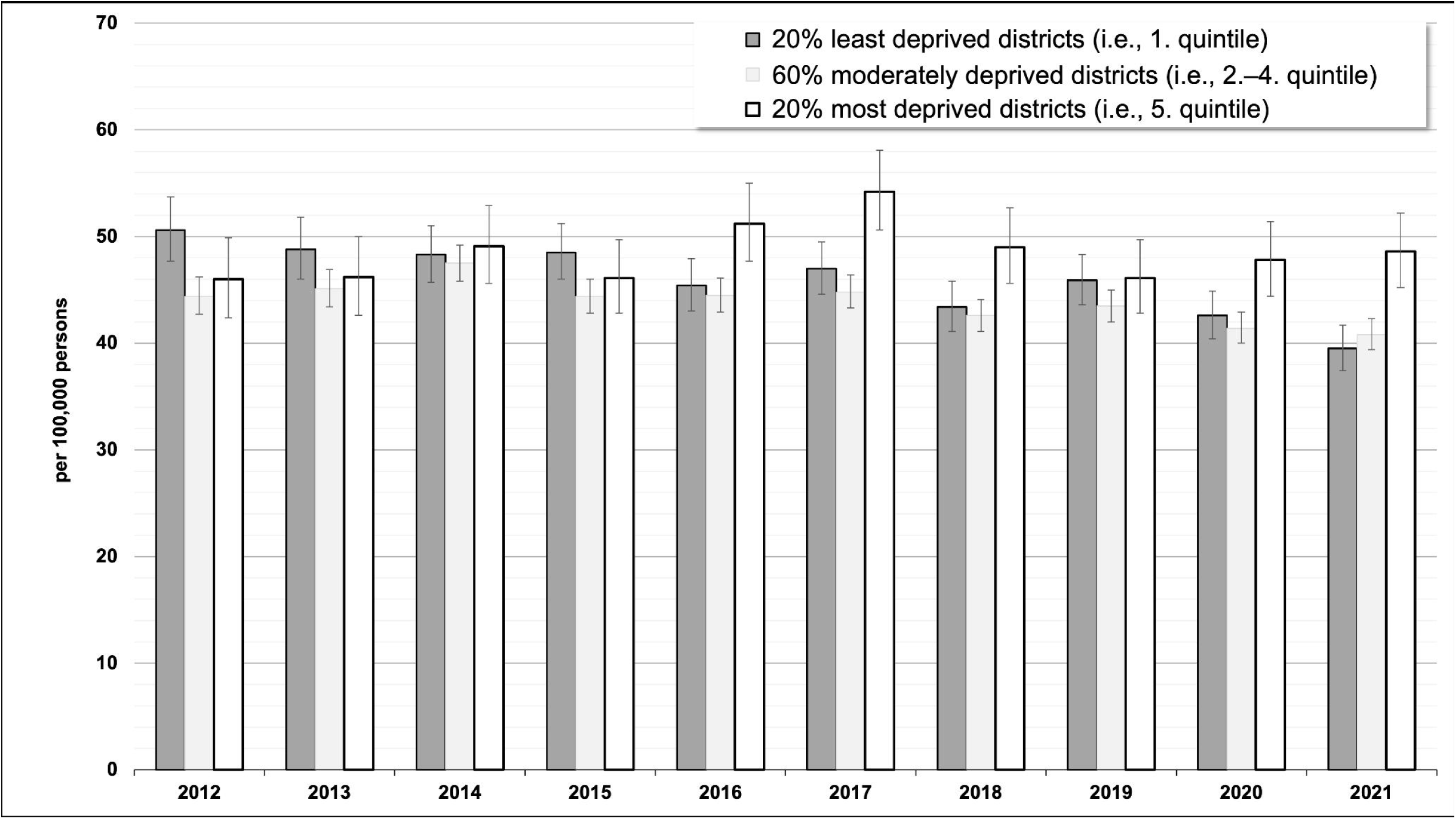
Standardized incidence proportions (with 95% CIs) of schizophrenia by district-level socioeconomic deprivation and calendar year

