## Supplementary Material for "Prevalence and incidence of schizophrenia: Temporal and regional trends in Germany"

**Table S1:** Antipsychotics used in this study for the identification of persons with treated schizophrenia

| **Drug name** | **ATC code** | **Antipsychotic class** |
| --- | --- | --- |
| Chlorpromazine | N05AA01 | Typical |
| Triflupromazine | N05AA05 | Typical |
| Cyamemazine | N05AA06 | Typical |
| Fluphenazine | N05AB02 | Typical |
| Perphenazine | N05AB03 | Typical |
| Thiopropazate | N05AB05 | Typical |
| Perazine | N05AB10 | Typical |
| Thioridazine | N05AC02 | Typical |
| Haloperidol | N05AD01 | Typical |
| Trifluperidol | N05AD02 | Typical |
| Bromperidol | N05AD06 | Typical |
| Benperidol | N05AD07 | Typical |
| Sertindole | N05AE03 | Atypical |
| Ziprasidone | N05AE04 | Atypical |
| Lurasidone | N05AE05 | Atypical |
| Flupentixol | N05AF01 | Typical |
| Zuclopenthixol | N05AF05 | Typical |
| Fluspirilene | N05AG01 | Typical |
| Pimozide | N05AG02 | Typical |
| Clozapine | N05AH02 | Atypical |
| Olanzapine | N05AH03 | Atypical |
| Quetiapine | N05AH04 | Atypical |
| Asenapine | N05AH05 | Atypical |
| Olanzapine and samidorphan | N05AH53 | Atypical |
| Sulpiride | N05AL01 | Atypical |
| Amisulpride | N05AL05 | Atypical |
| Risperidone | N05AX08 | Atypical |
| Zotepine | N05AX11 | Atypical |
| Aripiprazole | N05AX12 | Atypical |
| Paliperidone | N05AX13 | Atypical |
| Iloperidone | N05AX14 | Atypical |
| Cariprazine | N05AX15 | Atypical |
| Brexpiprazole | N05AX16 | Atypical |

**Table S2:** Standardized incidence proportion (with 95% CIs) of schizophrenia by demographic characteristics and calendar year (per 100,000 persons)

|  | **2012** | **2013** | **2014** | **2015** | **2016** |
| --- | --- | --- | --- | --- | --- |
| **Total number of (database) population, 0–64 years, n** | 9,589,084 | 9,879,340 | 10,958,044 | 11,361,701 | 11,403,109 |
| **Overall** |  |  |  |  |  |
| Both sexes | 46.0 (44.6; 47.4) | 46.0 (44.7; 47.4) | 47.7 (46.4; 49.1) | 45.5 (44.3; 46.8) | 45.5 (44.3; 46.8) |
| Females | 42.3 (40.6; 44.1) | 41.9 (40.1; 43.7) | 43.0 (41.4; 44.8) | 40.8 (39.3; 42.5) | 39.9 (38.3; 41.5) |
| Males | 49.6 (47.5; 51.7) | 50.1 (48.0; 52.2) | 52.3 (50.3; 54.4) | 50.0 (48.1; 52.0) | 51.0 (49.1; 53.0) |
| **Age group 0–17 years** |  |  |  |  |  |
| Both sexes | 4.1 (3.4; 5.1) | 3.5 (2.9; 4.4) | 3.7 (3.0; 4.5) | 3.6 (2.9; 4.4) | 4.1 (3.4; 5.0) |
| Females | 3.7 (2.7; 5.0) | 3.6 (2.7; 4.9) | 3.6 (2.7; 4.9) | 3.9 (2.9; 5.1) | 3.8 (2.9; 5.1) |
| Males | 4.6 (3.5; 6.0) | 3.5 (2.6; 4.7) | 3.7 (2.8; 4.9) | 3.3 (2.5; 4.4) | 4.4 (3.4; 5.7) |
| **Age group 18–64 years** |  |  |  |  |  |
| Both sexes | 57.4 (55.7; 59.2) | 57.6 (55.9; 59.4) | 59.7 (58.1; 61.4) | 56.9 (55.4; 58.5) | 56.8 (55.3; 58.4) |
| Females | 52.7 (50.5; 55.0) | 52.1 (50.0; 54.4) | 53.6 (51.5; 55.8) | 50.8 (48.8; 52.8) | 49.5 (47.6; 51.6) |
| Males | 62.0 (59.3; 64.7) | 63.0 (60.3; 65.7) | 65.7 (63.2; 68.3) | 62.9 (60.5; 65.4) | 63.9 (61.5; 66.4) |
| (1 of 2) | | | | | |
| Estimates on incidence proportion of schizophrenia are age- and sex-standardized to the population of Germany on December 31, 2021. | | | | | |

**Table S2 (continued):** Standardized incidence proportion (with 95% CIs) of schizophrenia by demographic characteristics and calendar year (per 100,000 persons)

|  | **2017** | **2018** | **2019** | **2020** | **2021** |
| --- | --- | --- | --- | --- | --- |
| **Total number of (database) population, 0–64 years, n** | 11,615,083 | 11,846,787 | 12,155,616 | 12,349,910 | 12,450,531 |
| **Overall** |  |  |  |  |  |
| Both sexes | 46.5 (45.2; 47.7) | 43.5 (42.3; 44.7) | 44.4 (43.2; 45.6) | 42.4 (41.3; 43.6) | 41.3 (40.1; 42.4) |
| Females | 41.8 (40.2; 43.5) | 38.2 (36.7; 39.7) | 38.3 (36.8; 39.8) | 37.7 (36.3; 39.2) | 37.1 (35.6; 38.6) |
| Males | 51.0 (49.1; 52.9) | 48.6 (46.8; 50.5) | 50.4 (48.6; 52.2) | 46.9 (45.2; 48.7) | 45.3 (43.6; 47.0) |
| **Age group 0–17 years** |  |  |  |  |  |
| Both sexes | 3.9 (3.2; 4.7) | 4.1 (3.4; 5.0) | 4.1 (3.4; 5.0) | 4.0 (3.3; 4.9) | 3.8 (3.1; 4.7) |
| Females | 3.9 (3.0; 5.2) | 3.3 (2.4; 4.5) | 3.9 (3.0; 5.2) | 4.2 (3.2; 5.6) | 4.4 (3.3; 5.7) |
| Males | 3.8 (2.9; 5.0) | 4.9 (3.8; 6.3) | 4.2 (3.2; 5.5) | 3.8 (2.9; 5.1) | 3.3 (2.4; 4.5) |
| **Age group 18–64 years** |  |  |  |  |  |
| Both sexes | 58.1 (56.5; 59.7) | 54.2 (52.7; 55.7) | 55.4 (53.9; 56.9) | 52.8 (51.4; 54.3) | 51.5 (50.0; 52.9) |
| Females | 52.0 (50.0; 54.0) | 47.6 (45.7; 49.5) | 47.5 (45.6; 49.4) | 46.7 (44.9; 48.6) | 45.9 (44.0; 47.7) |
| Males | 64.0 (61.6; 66.4) | 60.7 (58.4; 63.0) | 63.1 (60.8; 65.4) | 58.8 (56.6; 61.0) | 56.9 (54.8; 59.1) |
| (2 of 2) | | | | | |
| Estimates on incidence proportion of schizophrenia are age- and sex-standardized to the population of Germany on December 31, 2021. | | | | | |

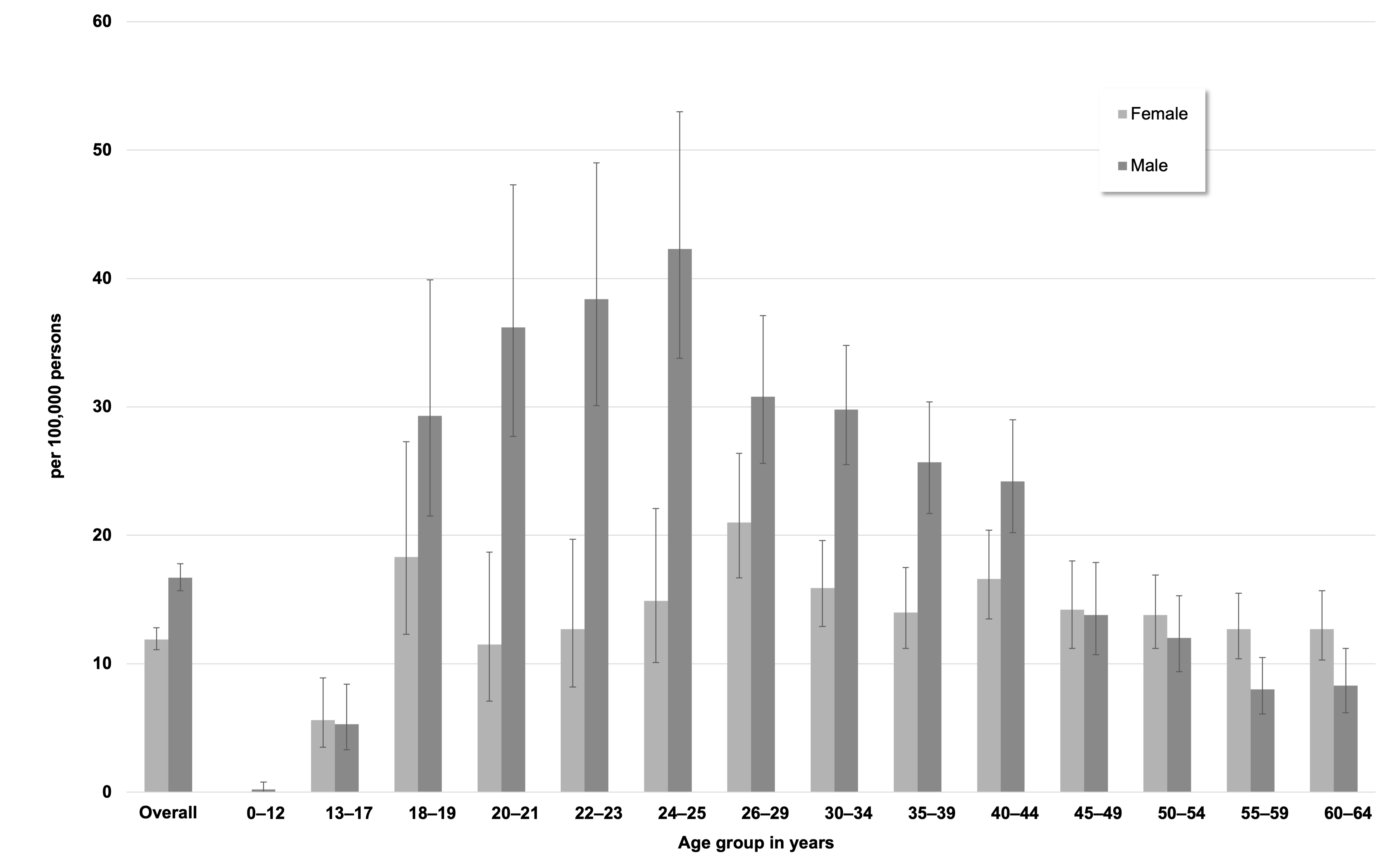

**Figure S1:** Standardized incidence proportions of schizophrenia (with 95% CIs) by sex and age in 2021 when considering only inpatient diagnoses

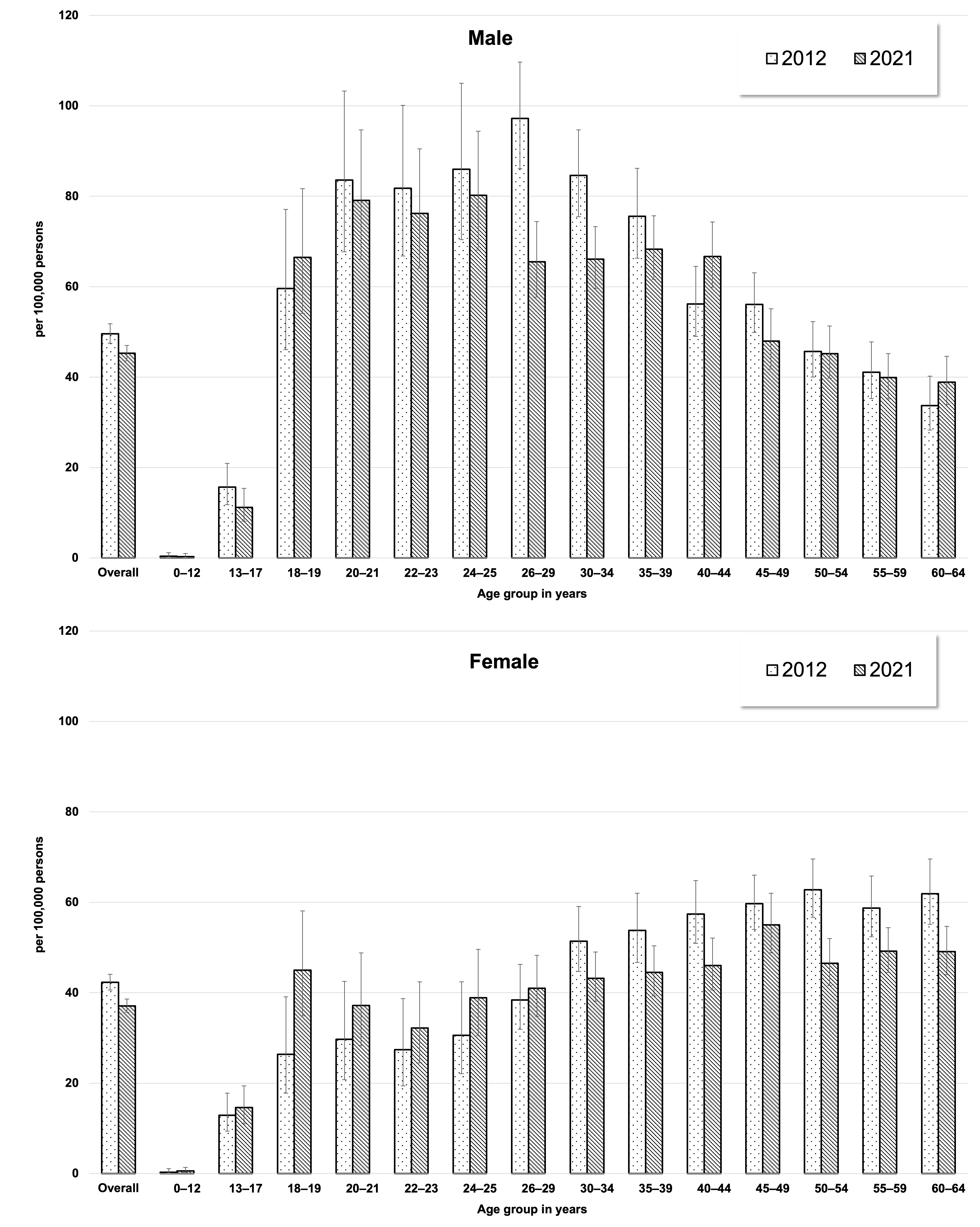

**Figure S2:** Standardized incidence proportions (with 95% CIs) of schizophrenia in 2012 and 2021 by age among males (upper figure) and females (lower figure)

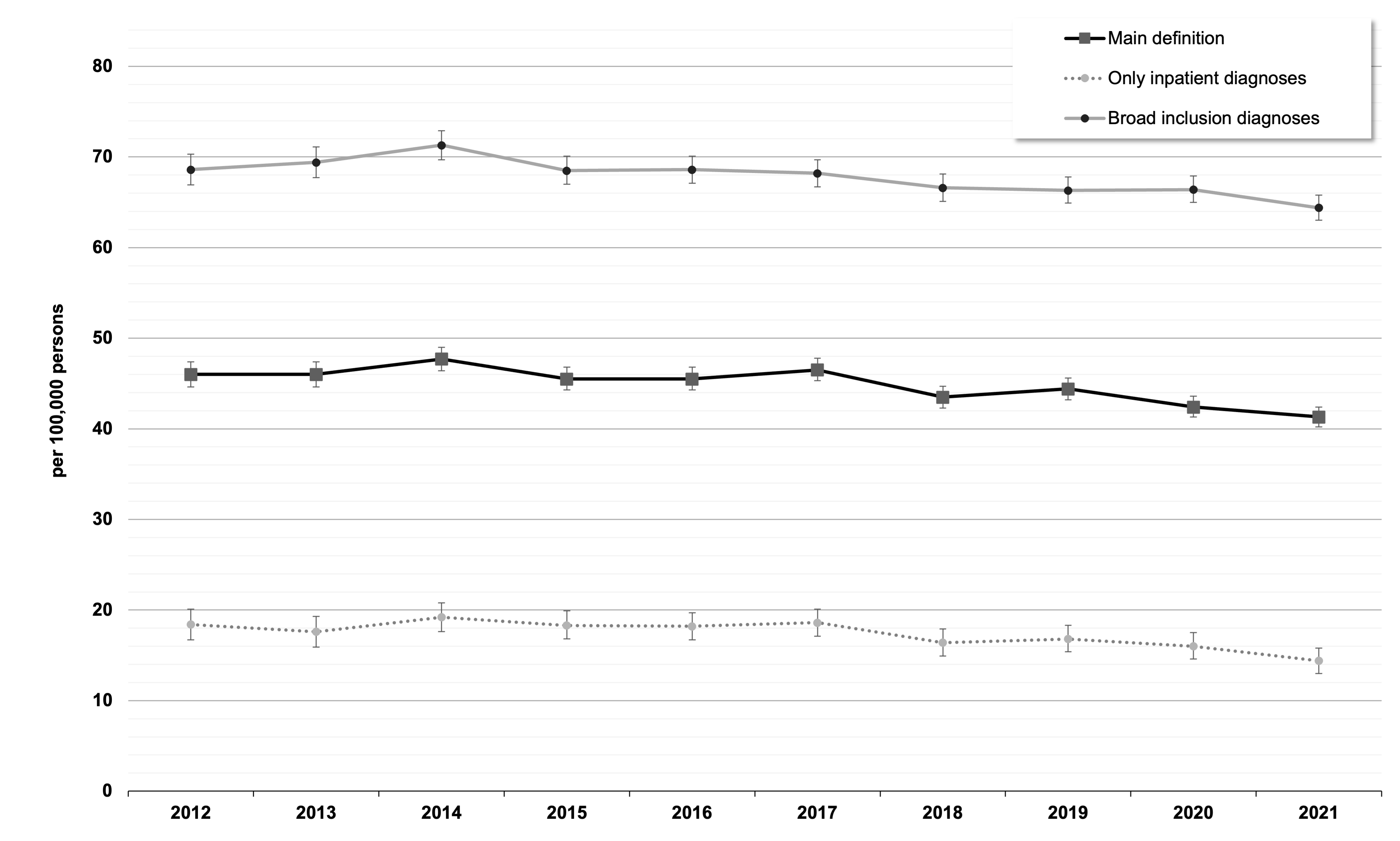

**Figure S3:** Overall standardized incidence proportion of schizophrenia (with 95% CIs) by calendar year according to (a) the main definition (antipsychotic plus outpatient or inpatient diagnosis F20), (b) when considering only inpatient diagnoses, or (c) when considering broad inclusion diagnoses (F20, F21, F22, F23, F25, F28, F29)

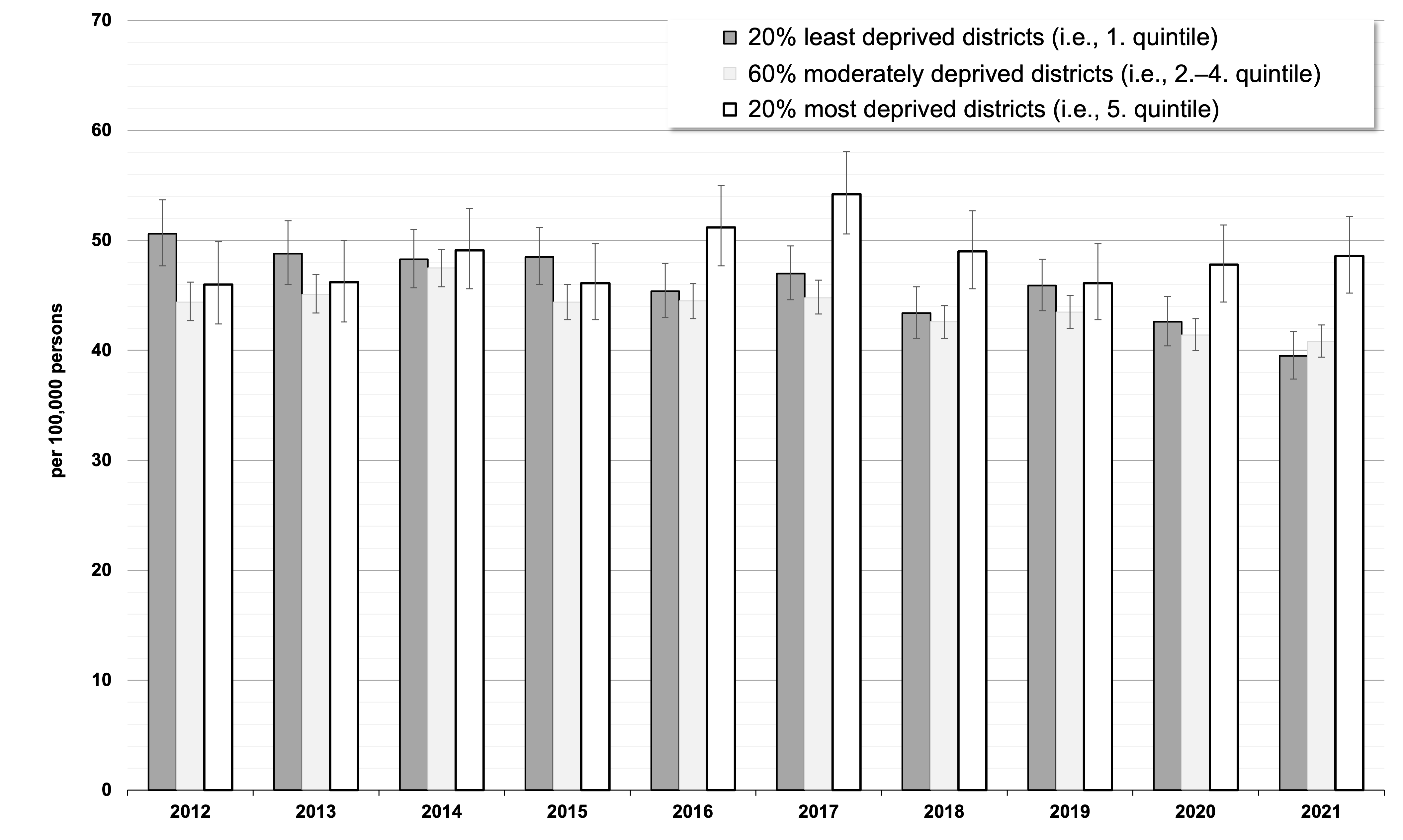

**Figure S4:** Standardized incidence proportions (with 95% CIs) of schizophrenia by district-level socioeconomic deprivation and calendar year
